# Validating the RISE UP score for predicting prognosis in patients with COVID-19 in the emergency department, a retrospective study

**DOI:** 10.1101/2020.11.23.20236786

**Authors:** Paul M.E.L. van Dam, Noortje Zelis, Patricia M. Stassen, Daan J.L. van Twist, Peter W. de Leeuw, Sander M.J. van Kuijk, Jacqueline Buijs

## Abstract

**Objective:** To mitigate the burden of COVID-19 on the healthcare system, information on the prognosis of the disease is needed. The recently developed RISE UP score has very good discriminatory value with respect to short-term mortality in older patients in the emergency department (ED). It consists of six items: age, abnormal vital signs, albumin, blood urea nitrogen (BUN), lactate dehydrogenase (LDH), and bilirubin. We hypothesized that the RISE UP score could have discriminatory value with regard to 30-day mortality in ED patients with COVID-19.

**Setting:** Two EDs of the Zuyderland Medical Centre (MC), secondary care hospital in the Netherlands.

**Participants:** The study sample consisted of 642 adult ED patients diagnosed with COVID-19 between March 3^rd^ until May 25^th^ 2020. Inclusion criteria were: 1) admission to the hospital with symptoms suggestive of COVID-19, and 2) positive result of the polymerase chain reaction (PCR), or (very) high suspicion of COVID-19 according to the chest computed tomography (CT) scan.

**Outcome:** Primary outcome was 30-day mortality, secondary outcome was a composite of 30-day mortality and admission to intensive care unit (ICU).

**Results:** Within 30 days after presentation, 167 patients (26.0%) died and 102 patients (15.9%) were admitted to ICU. The RISE UP score showed good discriminatory value with respect to 30-day mortality (AUC 0.77, 95% CI 0.73-0.81), and to the composite outcome (AUC 0.72, 95% CI 0.68-0.76). Patients with RISE UP scores below 10% (121 patients) had favourable outcome (0% mortality and 5% ICU admissions). Patients with a RISE UP score above 30% (221 patients) were at high risk of adverse outcome (46.6% mortality and 19% ICU admissions).

**Conclusion:** The RISE UP score is an accurate prognostic model for adverse outcome in ED patients with COVID-19. It can be used to identify patients at risk of short-term adverse outcome, and may help guiding decision-making and allocating healthcare resources.

## Background

The COVID-19 pandemic has affected millions of people, and has put enormous strains on healthcare resources.^[1-4]^ To mitigate the burden on the healthcare system, information on the prognosis of the patients is needed. Although most patients with COVID-19 develop only mild symptoms, some develop severe and potentially fatal complications.^[1, 2, 5, 6]^

Prediction models could help to estimate the risk of a poor or good outcome and may therefore assist in triaging patients when allocating healthcare resources. Many risk prediction models have been developed for patients with COVID-19, most of which still need rigorous external validation efforts to assess their added value to clinical practice. However, our group developed and validated the RISE UP score (Risk Stratification in the Emergency Department in Acutely Ill Older Patients), an accurate and easy to use model, predicting mortality in medical patients older than 65 years in the emergency department (ED).^[7]^ The model had very good discriminatory value with an area under the receiver operating characteristic curve (AUC) of 0.84 in the derivation cohort and 0.83 in the validation cohort.

We hypothesized that the RISE UP score may also be valuable in predicting prognosis in patients with COVID-19, as this disease predominantly affects the same age group. The aim of the present retrospective study was to evaluate the discriminatory value of the RISE UP score with regard to poor outcome (mortality and/or admission to the intensive care unit (ICU)) in ED patients with COVID-19.

## Materials and methods

This study has been reported in accordance with the Transparent reporting of a multivariable prediction model for individual prognosis or diagnosis (TRIPOD): The TRIPOD statement.^[8]^

### Study design and setting

We performed a retrospective analysis at the two EDs of the Zuyderland Medical Centre (MC), location Heerlen and Sittard-Geleen. This is one of the largest teaching hospitals in the Netherlands providing secondary care to a total of 55,000 patients during ED visits on a yearly basis. This study was approved by the medical ethics committee of the Zuyderland MC (METCZ20200136). Informed consent was obtained retrospectively by an opt-out method by writing a letter to the included patients.

### Study sample

The study sample consisted of consecutive adult (18 years or older) medical ED patients diagnosed with COVID-19 in the period from March 3^rd^ until May 25^th^ 2020. Patients were included if they met the following criteria: 1) admission to the hospital with symptoms suggestive of COVID-19 (i.e. coughing, common cold, sore throat, dyspnoea, acute diarrhoea, vomiting, fever, or an unexpectedly discovered oxygen saturation below 92%); and 2) positive result of the polymerase chain reaction (PCR) for SARS-CoV-2 in respiratory specimens; or (very) high suspicion of COVID-19 according to the computed tomography (CT) scan of the thorax (CO-RADS 4 or CO-RADS 5, see Supplementary Table).^[9]^ We excluded patients who revisited the ED after an earlier ED presentation during the study period.

**Supplementary Table.** Overview of CO-RADS categories for suspicion of pulmonary involvement of COVID-19^[9]^.

|  | Level of suspicion | Summary |
| --- | --- | --- |
| CO-RADS 1 | Very low | Normal or non-infectious |
| CO-RADS 2 | Low | Typical for other infection but not COVID-19 |
| CO-RADS 3 | Equivocal / unsure | Features compatible with COVID-19, but also other diseases |
| CO-RADS 4 | High | Suspicious for COVID-19 |
| CO-RADS 5 | Very high | Typical for COVID-19 |

CO-RADS, COVID-19 Reporting And Data System.

### Data collection

From electronic medical records, we collected data on age, sex, and information regarding comorbidity according to the Charlson Comorbidity Index (CCI).^[10]^ The following vital signs were retrieved: heart rate (HR), mean arterial blood pressure (MAP), respiratory rate (RR), oxygen saturation, temperature, and Glasgow Coma Scale (GCS). Supplemental oxygen was taken into account when measuring oxygen saturation. When the RR or GCS were missing, we used paCO2 and descriptions in the medical records to deduce these values, similar to other studies.^[7, 11, 12]^ In addition, we collected routinely assessed laboratory tests: blood urea nitrogen (BUN), lactate dehydrogenase (LDH), bilirubin, albumin, lymphocytes, and C-reactive protein (CRP).

We retrieved all items of the RISE UP score. The original formula of the RISE UP score to calculate an individual’s probability of 30-day mortality is: P(30-day mortality) = 1 / (1 + exp (-(−3.908 + 0.050*Age + 1.115*≥2 Abnormal Vital Signs (yes=1, no=0) – 0.112*Albumin (in g/L) + 0.284*(BUN (in mmol/L)/5) + 0.120*(LDH (in U/L)/100) + 0.875*Bilirubin >20 µmol/L (yes=1, no=0))).^[7]^

Furthermore, we collected the results of the reverse-transcriptase PCR for SARS-CoV-2 in respiratory specimens and the results of the CT scan of the thorax.^[9]^ Finally, we retrieved data on length of hospital stay, admission to the intensive care unit (ICU) and mortality. Data on mortality were verified using medical records, which are connected to the municipal administration office.

### Outcomes

The primary outcome was all-cause mortality within 30 days after ED presentation. The secondary outcome was a composite of mortality within 30 days after ED presentation and admission to the ICU. In our hospital, the main reason for admitting patients to the ICU was mechanical ventilator support.

### Statistical analysis

Baseline characteristics were analysed using descriptive statistics on the observed data. For each included patient, we calculated their individual probability of 30-day mortality and of the composite endpoint of 30-day mortality and/or admission to ICU using the initial RISE UP score formula. In case the probability could not be computed in over 5% of patients due to missing values, data would be imputed using multiple imputation, as per our study protocol. Predictive performance of the RISE UP score in this external validation was quantified as discriminative ability (i.e., the ability of the model to separate those who will experience the event from those who will not) and model calibration. We calculated the AUC of the receiver operating characteristic (ROC) curve to quantify the discriminative ability. An AUC of 0.5 would correspond to no more discriminative value than the toss of a coin, whereas an AUC of 1.0 would mean perfect separation. Calibration of the RISE UP score was assessed by visually inspecting the calibration plot that shows the agreement between predicted and observed probabilities for subgroups of patients based on similar predicted risk. In case of poor calibration (i.e., the intercept of the plot is not equal to 0 and/or the slope of the plot is not equal to 1), the model was recalibrated using logistic regression analysis with the linear predictor of the RISE UP score, the linear sum of the regression coefficient multiplied by their respective predictor values, as the only independent variable.

Finally, we aimed to identify cut off values of the RISE UP score that will support clinical decision-making, by identifying patients with low and high risk of an adverse outcome (low risk and high risk group). To achieve this, we calculated sensitivity, specificity, positive and negative predictive values from multiple cut off values of the RISE UP score.

All data were analysed using IBM SPPS Statistics for Windows, IBM Corporation, Armonk N.Y., USA, version 25.0.

### Patient and public involvement

The patients and the public were not involved in the design, conduct, reporting, or dissemination plans of our research.

## Results

### Study sample

During the study period, 642 consecutive ED patients diagnosed with COVID-19 met the inclusion criteria and were included (Table 2). The median age of patients was 72 years (IQR 62-80), and 69.2% were 65 years or older. Most patients (63.4%) were male. More than half of the patients (59.8%) had two or more abnormal vital signs. The laboratory results showed elevated LDH (median 350 U/L). The median length of hospital stay was 6 days (IQR 3-12).

**Table 2.** Baseline characteristics of study population.

|  | Reference values | Study sample (n = 642) |
| --- | --- | --- |
| Age |  |  |
| Age, median (IQR), years |  | 72 (62-80) |
| 65 years or older, n% |  | 444 (69.2) |
| Male, n% |  | 407 (63.4) |
| CCI score, median (IQR) |  | 1 (0-3) |
| Abnormal vital signs, n% |  |  |
| Tachycardia (HR > 90 bpm) |  | 330 (51.4) |
| Hypotension (MAP < 70 mmHg) |  | 13 (2.0) |
| Tachypnoe (RR > 20/min) |  | 367 (57.2) |
| O2 saturation < 95% |  | 325 (50.6) |
| Hypo- or hyperthermia (< 36 or > 38 °C) |  | 332 (51.7) |
| GCS < 15 |  | 96 (15.0) |
| ≥ 2 abnormal vital signs <sup>a</sup> |  | 384 (59.8) |
| Laboratory results |  |  |
| Lymphocytes, median (IQR), x 10<sup>9</sup>/L | 1.1 – 4.0 | 0.85 (0.59-1.22) |
| BUN, median (IQR), mmol/L | 3.5 – 7.5 | 6.9 (5.0-10.0) |
| Bilirubin, median (IQR), µmol/L | < 21 | 9.0 (7.0-13.0) |
| LDH, median (IQR), U/L | < 248 | 350 (269-462) |
| Albumin, mean (SD), g/L | 35 – 50 | 37.4 (4.3) |
| CRP, median (IQR), mg/L | < 10 | 77 (36-130) |
| Positive PCR for SARS-CoV-2, n% |  | 553 (86.1) |
| CO-RADS 4 or CO-RADS 5 on CT thorax, n% |  | 366 (57.0) |
| Intensive care unit (ICU) admission, n% |  | 102 (15.9) |
| Length of hospital stay, median (IQR), days |  | 6 (3-12) |
| Outcome |  |  |
| Mortality within 30 days, n% |  | 167 (26.0) |
| Composite endpoint (mortality and/or ICU), n% |  | 226 (35.2) |
| Number of days until death, median (IQR) |  | 8 (4-15) |

BUN, blood urea nitrogen; CCI, Charlson comorbidity index; CO-RADS, COVID-19 Reporting And Data System; CRP, C-reactive protein; GCS, Glasgow coma scale; HR, heart rate; IQR, interquartile range; LDH, lactate dehydrogenase; MAP, mean arterial pressure; PCR, polymerase chain reaction; RR, respiratory rate; SD, standard deviation.
<sup>a</sup> ≥ 2 of following abnormal vital signs: HR > 90 bpm, MAP < 70 mmHg, RR > 20/min, O2 saturation < 95% and GCS < 15.

In our sample, 102 patients (15.9%) were admitted to ICU and 167 patients died, yielding a 30-day mortality of 26.0%. The median number of days until death was 8 (IQR 4-15).

### RISE UP score

The RISE UP score could not be calculated in 21 patients (3.2%) because of missing values (laboratory tests). As this was below our 5% threshold, no imputation step was performed. For 621 patients the RISE UP score yielded an AUC of 0.77 (95% CI: 0.73-0.81) with respect to the 30-day mortality, and an AUC of 0.72 (95% CI 0.68-0.76) with respect to the composite endpoint of 30-day mortality and/or admission to the ICU.

The initial calibration plot showed average underestimation of 30-day mortality by the model and a slope of >1. This could at least in part be due to the fact that in the derivation and validation cohort of the RISE UP score we observed a 30-day mortality of 10.9% and 13.3%, respectively. Therefore, we performed recalibration of the model.^[7]^ The formula was adjusted by changing the intercept to - 2.083 and multiplying the linear sum of regression coefficients multiplied by their respective predictor values by 0.795. The adjusted formula for the RISE UP score in patients with COVID-19 was: P(30-day mortality) = 1 / (1 + exp (−(−2.083 + 0.795 * (0.050*Age + 1.115*≥2 Abnormal Vital Signs (yes=1, no=0) – 0.112*Albumin (in g/L) + 0.284*(BUN (in mmol/L)/5) + 0.120*(LDH (in U/L)/100) + 0.875*Bilirubin >20 µmol/L (yes=1, no=0)))).

### Determination of clinical cut off values

The individual RISE UP scores were stratified according to their probability to predict 30-day mortality. Patients with a RISE UP score <10% were considered to be at (very) low risk of mortality (Table 3). In this subgroup of 121 patients (19.5%), there were no deaths and only six (5.0%) were admitted to ICU, yielding a sensitivity of 100% (95% CI 97.7-100) and a negative predictive value of 100%. A RISE UP score <5% also yielded a sensitivity and negative predictive value of 100%, however, this subgroup of patients consisted of 41 patients only (6.6%).

**Table 3.** Cut off values of the RISE UP score.

| Cut off value (%) | Patients (n%) <sup>a</sup> | Deaths (n%) <sup>b</sup> | Admissions to ICU (n%) <sup>b</sup> | Sensitivity (% , 95% CI) <sup>c</sup> | Specificity (% , 95% CI) <sup>c</sup> | PPV (% , 95%CI) <sup>c</sup> | NPV (% , 95% CI) <sup>c</sup> |
| --- | --- | --- | --- | --- | --- | --- | --- |
| <5 | 41 (6.6) | 0 (0) | 3 (7.3) | 100<br>(97.7-100) | 8.9<br>(6.5-11.9) | 27.8<br>(27.3-28.4) | 100<br>(97.7-100) |
| <10 | 121 (19.5) | 0 (0) | 6 (5.0) | 100<br>(97.7-100) | 26.3<br>(22.3-30.6) | 32.2<br>(31.1-33.5) | 100<br>(97.7-100) |
| >30 | 221 (35.6) | 103 (46.6) | 42 (19.0) | 64.0<br>(56.1-71.4) | 74.3<br>(70.1-78.3) | 47.4<br>(41.9-51.6) | 85.5<br>(82.6-87.4) |
| >50 | 74 (11.9) | 35 (47.2) | 11 (14.8) | 21.7<br>(15.6-28.9) | 91.5<br>(88.6-93.9) | 47.3<br>(37.2-57.8) | 77.0<br>(75.3-78.4) |

CI, confidence interval; ICU, intensive care unit; n, number of cases; NPV, negative predictive value; PPV, positive predictive value; RISE UP, Risk stratification in the emergency department in acutely ill older patients.
<sup>a</sup> The total number of patients described in this table is 621, because of missing the RISE UP scores in 21 patients.
<sup>b</sup> The percentages of deaths and ICU admissions are related to the amount of patients above or below that cut off value.
<sup>c</sup> Sensitivity, specificity, PPV and NPV are calculated for mortality only.

Patients with a RISE UP score >30% were at high risk of a poor outcome (Table 3). In this subgroup of 221 patients (35.6%), we found a mortality of approximately 50% and 15% ICU admission.

## Discussion

In this retrospective study in two EDs in the Netherlands, we found that the RISE UP score showed good discriminatory value with respect to 30-day all-cause mortality and/or admission to ICU in ED patients with COVID-19. The model yielded an AUC of 0.77 (95% CI 0.73-0.81) for 30-day mortality and an AUC of 0.72 (95% CI 0.68-0.76) for a composite of 30-day mortality and/or admission to ICU. After performing recalibration the RISE UP score was well calibrated. We found that patients with a RISE UP score <10% had favourable outcomes and that patients with a RISE UP score >30% were at high risk of adverse outcomes.

Using the RISE UP score, the probability of both favourable and poor outcomes can be predicted in the first two hours of the ED visit. This is important because this may guide clinical decision-making, especially during the current COVID-19 pandemic with healthcare system facing problems with clinical healthcare facilities (i.e. shortage of ICU beds) and the development of out-of-hospital low care facilities (i.e. corona hotels). The finding that the RISE UP score may be useful to guide clinical decision-making is important, because a clinical algorithm like this score is essential for allocation of healthcare resources. The RISE UP score is applicable for both sides of the clinical spectrum. In our cohort, 121 patients (19.5%) had a RISE UP score <10%, and had very low risk of short-term mortality or ICU admission. In these patients, the clinician can choose safely to refer the patient to outpatient treatment or to discharge at an early stage. On the other hand, the 74 patients (11.9%) in our cohort with a RISE UP score >30% had a high risk of adverse outcome. This may support the decision to transfer these patients to ICU at an early stage. Furthermore, in patients with high RISE UP scores, pre-existing multiple comorbidities and poor clinical performance, the model might support the decision to choose supportive care only.

The RISE UP score was recently developed to predict 30-day all-cause mortality in older medical ED patients. The score consists of six easily and readily available items during the ED visit.^[7]^ The RISE UP score can be easily implemented in routine practice with an online calculator (https://jscalc.io/calc/o1vzp36bIDGQUCYl). The model has face validity as it includes items that are markers of severe disease, such as sepsis, renal failure, liver failure, or shock. Because of this, the RISE UP score reflects the severity of illness in ED patients. Therefore, it is not surprising that the model has good discriminatory value for poor outcome in ED patients with COVID-19, since our sample largely consisted of older medical patients (69.2% were 65 years or older). Other studies have also shown higher mortality in older patients with COVID-19.^[13-16]^ We state that application of the RISE UP score easily identifies patients with COVID-19 who are at high and at low risk of dying within the first 30 days after presentation to the ED.

The original research article concerning the RISE UP score showed a better predictive performance compared to our current study (AUC 0.83 versus 0.77).^[7]^ The difference can be explained in several ways. First, 30-day mortality was around 11% in the original cohort, while in our cohort it was 26.0%.^[7]^ This can be explained because in this study, we only included ED patients who were admitted to the hospital, whereas patients that were discharged from the ED were excluded. Furthermore, COVID-19 can have a serious course in admitted patients. Therefore, the probability of a poor outcome was higher in our cohort. Secondly, the levels of LDH were often elevated in patients with COVID-19 (81.1% were higher than the reference value). Since LDH is one of the items in the RISE UP score, this may affect the discriminatory value. Third, in our clinical experience, some patients with COVID-19 show a rapid deterioration of their clinical condition, while there were few objective symptoms in the initial assessment. This phenomenon may play a role as well, since the RISE UP score includes the initial assessment only.

The RISE UP score still has good discriminatory value in a population that is different from the population in which it was designed. This can be explained because the model accurately reflects the severity of illness in ED patients. The severity is determined by both the severity of the disease, and pre-existing comorbidities. In COVID-19, the prognosis is also determined by the disease itself and patient related factors (i.e. age and comorbidities).^[1, 5]^

In a recent systematic review, a total of 16 prognostic models for predicting mortality, progression to severe disease or length of hospital stay in patients with COVID-19 were analysed.^[17]^ The AUC estimates ranged from 0.85 to 0.99. However, most proposed models were not clearly described and therefore it is unclear whether they are feasible in other settings. The authors of the systematic review do not recommend the use of any of these models in current practice.^[17]^ New prediction models will be developed for patients with COVID-19, such as two recently published studies, that show good results and should definitely be included in future studies.^[18, 19]^ The RISE UP score is an accurate and externally validated prediction model with good discriminatory value in unselected older medical patients.^[7]^ We state that the RISE UP score can be used in current clinical practice.

Our study has several limitations. First, our study was performed in the two EDs of one medical centre which may limit generalizability of the results. However, our study has a relatively large cohort of patients with COVID-19 in one of the most heavily affected areas of the Netherlands. Second, 89 patients (13.9%) did not have a positive PCR result. Because there is no gold standard for the diagnosis of COVID-19 and the diagnostic accuracy of the PCR is limited, we believe it is diligent to also include patients with symptoms suggestive of COVID-19 and a positive result of the chest CT scan. Other studies have used these inclusion criteria as well.^[20]^ If we apply the RISE UP score only to the patients that were confirmed with PCR, we found similar discriminatory values (AUC 0.76 (95% CI: 0.71-0.80) for 30-day mortality). Next, our study was limited to patients admitted to the hospital. It would be interesting to apply the RISE UP score to patients who would be discharged. However, these data were not available to us. Finally, the number of ICU admissions in our study was relatively low (15.9%), which may result from decisions to initiate conservative care in patients with pre-existing frailty or severe comorbidity. These decisions may be different in other countries, which made us decide to study ICU admissions as a composite outcome only. Since existing literature is dominated by predicting mortality in patients with COVID-19, we consider the inclusion of ICU admission a strength of our current study.

The RISE UP score can be readily implemented in routine practice at the ED. It is easy to use and not time-consuming to fill in. Our current study shows that the discriminatory value in ED patients with COVID-19 is good and that the RISE UP score is therefore also applicable in these patients.

## Conclusion

In conclusion, the RISE UP score is an accurate and easily available prediction model for 30-day mortality and ICU admission in ED patients with COVID-19. The score is useful to identify patients at low and high risk for clinical complications. So this model may contribute to fast recognition of patients who are at low or high risk for short term adverse outcome, and guide clinical decision-making and allocating healthcare resources in this severe pandemic, dealing with scarcity of clinical facilities and materials.

## Data Availability

All data referred to in the manuscript will be available upon reasonable request

## Contributors

PD, NZ, PMS, DT and JB collected the clinical data. PD, NZ and SMJK performed the statistical analysis. Data was interpreted by all authors. PD drafted the first version of the manuscript. NZ, PMS, PWL, SMJK, DT and JB critically reviewed the manuscript. All authors have read and approved the final version of the manuscript.

The corresponding author has the right to grant on behalf of all authors and does grant on behalf of all authors, an exclusive licence (on non-exclusive for government employees) on a worldwide basis to the BMJ Publishing Group Ltd to permit this article (if accepted) to be published in BMJ editions and any other BMJPGL product and sublicences such use and exploit all subsidiary right, as set in our licence. The corresponding author affirms that the manuscript is an honest, accurate, and transparent account of the study being reported. No important aspects of the study have been omitted.

---

**Figure 1.**
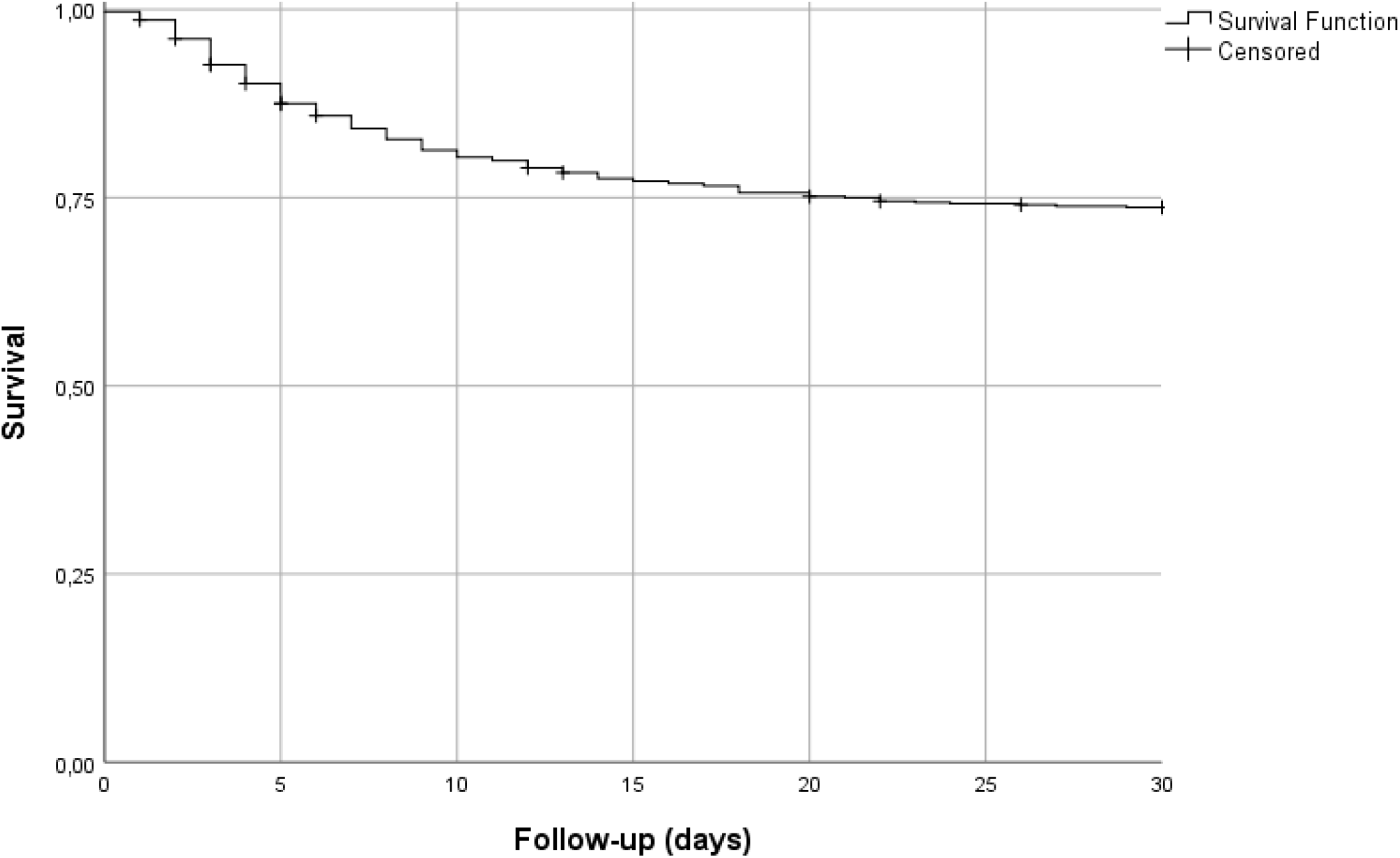
Kaplan Meier curve.

**Figure 2.**
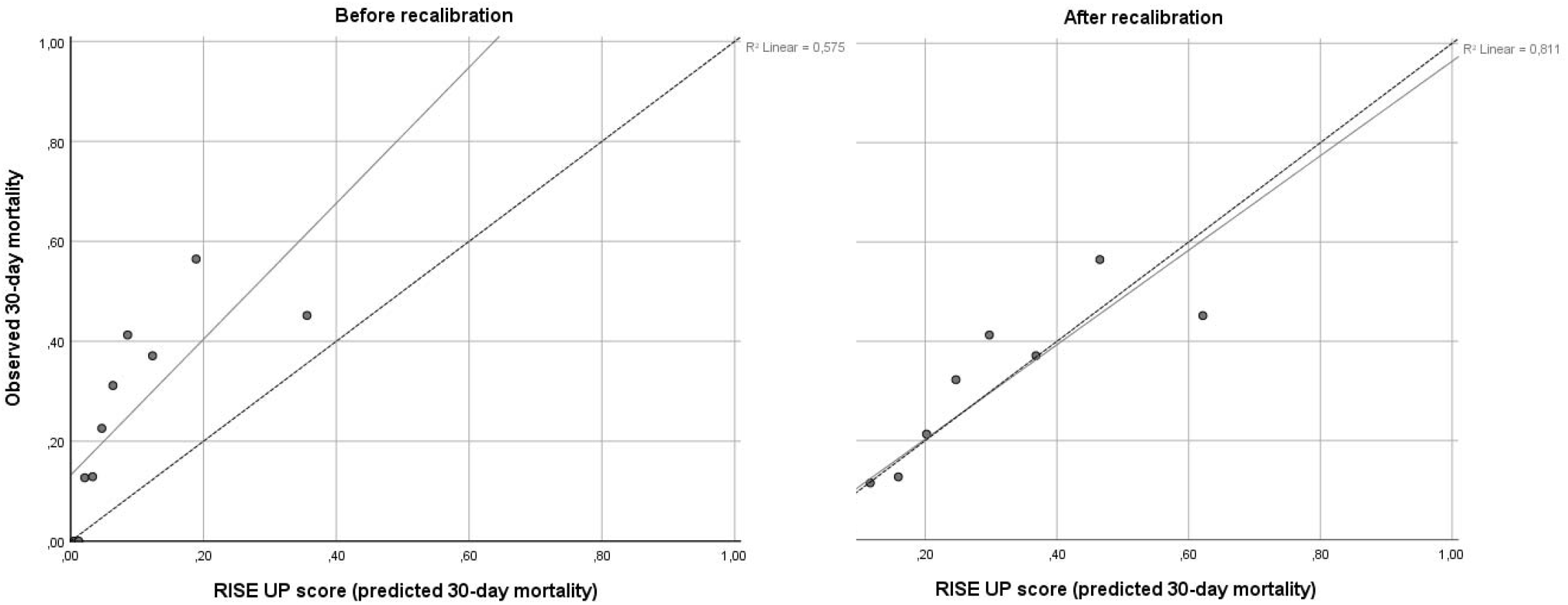
Calibration plots of the predicted 30-day mortality (x axis) versus the observed 30-day mortality (y axis) is represented by the diagonal line. The left calibration plot shows calibration of the RISE UP score before recalibration. The right calibration plot shows calibration of the RISE UP score after recalibration. The adjusted formula of the RISE UP score in COVID-19: P(30-day mortality) = 1 / (1 + exp (−(−2.083 + 0.795 * (0.050*Age + 1.115*≥2 Abnormal Vital Signs (yes=1, no=0) – 0.112*Albumin (in g/L) + 0.284*(BUN (in mmol/L)/5) + 0.120*(LDH (in U/L)/100) + 0.875*Bilirubin >20 µmol/L (yes=1, no=0)))).

